# Effect of COVID-19 on Food Choices and Eating Behaviour: a study protocol

**DOI:** 10.1101/2022.09.28.22280475

**Authors:** Jessica C. McCormack, Emily Doughty, Shaina Ebron, Mei Peng

## Abstract

**Background and Aims:** Research suggests that many individuals infected with COVID-19 experience changes in taste and smell that can persist for months after the initial infection. These sensory changes can potentially have long-term impacts on dietary choices, nutrition, and body weight. The aim of this study is to explore COVID-related changes in dietary intake among University Students.

**Methods:** A retrospective cohort design will be used to compare a cohort of University Students who experienced COVID-19 infection versus pre-existing data collected from a similar cohort prior to the pandemic. Specifically, the pre-existing data were collected between July 2017 and July 2021. Both datasets comprise of a weighed Food Record and Dutch Eating Behaviour Questionnaire. The cohort will also be asked about their experience with COVID-19 and changes in their eating behaviour since before the pandemic. Total daily energy intake, macronutrient intake and composition, will be compared across groups using an ANCOVA analysis controlling for age, gender, and ethnicity.

**Discussion:** Understanding the long-term impact of COVID-19 infection is crucial. While COVID-related sensory changes are hypothesised to have impacts on eating behaviour and dietary choices, it is challenging to perform controlled cohort studies due to the high prevalence of undetected infections. The proposed temporal analyses provide a unique opportunity to test for COVID-related impacts on eating behaviour.

## 1 Introduction

Research has suggested that many individuals who are infected with SARS-CoV-2, the virus responsible for the current COVID-19 pandemic, experience sensory symptoms, such as loss or distortion of smell or taste abilities. Sensory changes have a significant impact on preference and selection of foods, which can directly alter energy and macronutrient intake, as well as diet quality (1, 2). However, effects of COVID-19 on dietary choices are not yet clear. Importantly, the ability to conduct cross-sectional or case-control studies comparing exposed and unexposed individuals is hindered by the surge in COVID-19 infections since early 2022 in New Zealand, and limited access to PCR testing.

Our Sensory laboratory possesses datasets collected prior to the COVID-19 pandemic from experiments regarding eating behaviour and food choices amongst University student cohorts, providing key baseline data for the proposed comparisons. We therefore propose this study to conduct a natural experiment to assess possible changes in eating behaviour amongst university students.

### 1.1 New Zealand Exposure to COVID-19

Due to the public health measures put in early 2020, New Zealand had relatively low exposure to COVID-19 up until late 2021. On March 25, 2020 New Zealand entered a National lockdown that closed schools, workplaces, and all retail except supermarkets, pharmacies, and petrol stations and restricted entry into New Zealand from overseas (3). As a result of these measures New Zealand was able to eliminate spread of COVID-19 in the community and had a relatively low burden of disease (3, 4).

In December 2021 New Zealand moved form an elimination strategy to a minimisation and protection strategy, which implemented a vaccine and mask requirements for high-risk settings and activities (5). This was accompanied by loosening of border restrictions and quarantine requirements for vaccinated New Zealanders returning from overseas. Cases of COVID-19 infection in New Zealand surged from early February 2022, peaking at 22,025 new daily infections on 6 March 2022 (6).

The University of Otago, Dunedin is located in the South Island of New Zealand approximately 1430 kms from Auckland, which has been the New Zealand epicentre for most of the COVID-19 pandemic. At the University of Otago exposure to COVID-19 remained low for most of 2020 and 2021. In 2021 the total number of cases of COVID-19 for the Southern District Health Board (DHB) which covers a geographic region of 65,608 square km including Dunedin, was 234 cases (7). As of 14 September 2022 there have now been 129,853 cases of COVID-19 in the Southern DHB (8), suggesting almost 40% of the population of the Southern DHB population has been infected with COVID-19.

### 1.2 Sensory impact of COVID-19

Relatively early in the pandemic, changes in smell and taste were identified as distinctive symptoms of COVID-19 with potential diagnostic value (9, 10). Meta-analyses have found that sensory changes are highly prevalent, with as many as 40-60% of people reporting changes in smell or taste (11, 12), and with 20% of patients reporting onset of smell and taste changes prior to other symptoms (12). Studies also suggest, however, that the prevalence of sensory symptoms may vary by region (13) and may be less common in more recent COVID-19 variants (14). For example, only 11% of Australian COVID-19 patients reported sensory changes compared to 55% of European COVID-19 patients (13). A study comparing chemosensory dysfunction in a cohort of patients infected in January and February 2022 to those infected in March and April 2020 found that self-reported chemosensory dysfunction was significantly lower in those infected in 2022 (15); less than 25% of those infected in early 2022 reported altered sense of smell compared to more than 60% in early 2020. Similarly, an analysis of 12 reports of patients infected with COVID-19 during the Omicron wave had a pooled estimated of olfactory dysfunction of only 13% (14).

Although most patients recover their sense of smell and taste within three months of infection (16), studies have reported long-term impact of COVID-19 on smell and taste for some patients even after they have recovered from COVID-19. For example, a study recruiting patients with symptoms of smell distortion found 83% had long-term distortion of smell related to COVID-19 (17). Similarly, in a longitudinal survey of adults with COVID-19 and sudden change in smell or taste in early 2020, only 38.2% reported a complete recovery of their sense of smell or taste more than 2 years after initial infection (16). Long-term dysfunction of taste and smell can have a significant impact of quality of life, including reduced appetite and enjoyment of food, concern about body odours, and risk of injury (18).

### 1.3 Changes in Food Choice and Diet due to COVID-19

Changes in smell and taste perception due to COVID-19 infection have potential consequences for food choices and diet. While there is limited research of direct links between COVID-related sensory symptoms and eating behaviour, previous research in anosmia and hyposomnia had consistently indicated associated changes in eating habits, which in turn impact on nutrition and body weight (19).

A number of studies have identified changes in eating habits since the start of the COVID-19 pandemic, although findings are mixed and are not directly linked to COVID-19 infection. These changes include increases in snacking behaviours and comfort eating (20, 21), increased adherence to a healthy diet (22), increased consumption of high energy density foods (23–25), increased home cooking (22, 26), and increased alcohol consumption (27). Negative changes in dietary behaviour – such as a snacking and alcohol consumption – were associated with increased weight gain and reduced physical activity (20). In addition, recent research of children has highlighted increased weight issues during the pandemic (28).

There are a number of factors that have likely contributed to dietary changes over the last two years, including supply chain shortages, COVID-19 lockdowns, the Russian invasion of Ukraine, climate consciousness, and increasing food prices due to inflation. It is unclear whether the combination sensory changes and lockdown lifestyles during COVID-19 are responsible for long-term dietary changes. To date, there has not been any empirical study that specifically evaluates possible shifts in eating behaviour due to COVID-19 infection.

In this project, we propose to survey University students’ dietary behaviour, including food choices and intake, post COVID infection. Given the high transmissibility of the COVID-19 omicron variant and its mild symptoms, as well as changes to contact tracing and testing, it is likely that some people may have become infected with COVID-19 without being aware of their exposure, which makes it difficult to confirm individual exposure status. In order overcome these limitations we will compare participants who were infected with COVID-19 to pre-existing data collected by the Sensory Neuroscience Lab between July 2017 and July 2021 (i.e., prior to the pandemic and widespread infection) that used the same measures. Our laboratory possess data from previous experiments regarding eating behaviour and food choices amongst the University student cohort.

Specifically, we will test for difference in food choices, macro- and micro-nutrient compositions of diet, intake, and eating behaviour traits, across datasets collected before and after the pandemic.

### 1.4 Objective

The proposed study aims to test for difference in food choices, eating behaviour, dietary intake and composition amongst university students before and after the COVID-19 pandemic.

## 2 Methods

### 2.1 Study Design

A retrospective cohort design will be used to compare a cohort of University Students post COVID-19 infection and pre-existing cohort data collected before the COVID-19 pandemic. Current data will be collected via an online survey, in-person visit, and a paper food record.

The research has been approved by the University of Otago Ethics Committee (22/097).

### 2.2 Setting

Participants will be primarily drawn from the University student cohort at the University of Otago, Dunedin.

### 2.3 Participants

Participants will be eligible to participate if they meet the following criteria:

- Aged 18 or older
- University student
- Generally healthy (i.e., not on regular medication)
- Have been infected with COVID-19
- COVID-19 infection was confirmed at the time by PCR test or RAT
- Able to give e-consent

Participants meeting the following criteria will be excluded:

- Under 18 years of age
- Taking regular medication to manage a physical condition (not including contraceptive pill)
- Have a chronic sensory (taster or smell) dysfunction
- Have been diagnosed with an eating disorder in the last 2 years
- Currently on a weight-loss or weight gain dietary programme

### 2.4 Recruitment

Participants will be recruited through a combination of methods including social media, flyers, and word of mouth. All eligible participants will be entered into a prize draw to win up to NZ$100 in vouchers.

All advertisement will include a link to the participant information and e-consent form, and will automatically be sent a link to the online survey on submission of their consent form.

### 2.5 Pre-COVID dataset

Data were collected between July 2017 and July 2021 as part of research projects conducted at the University of Otago Sensory Science Lab. Participants provided consent for their data to be used in future research as part of obtaining their consent to participate in the research. Ethical approval for these studies was obtained from the University of Otago Ethics Committee.

Although this data comes from multiple studies, all studies drew from the same population of University of Otago students and the inclusion and exclusion criteria were consistent across studies. All studies include the same measures for eating behaviour and food intake, as well as assessment of height and weight.

### 2.6 Sample size

The sample size calculation was based on changes in energy intake in university students before and during the COVID-19 pandemic reported in previous studies (25, 29, 30). Based on a mean effect size of 0.22, in order to detect significant difference in energy intake (*p* < 0.05) with 80% power we will recruit a sample of 326 participants exposed to COVID-19 infection, for a total sample of approximately 920 participants.

### 2.7 Primary Outcome

The primary outcome is energy intake which will be assessed using a 24-hour Weighed Food Record. A 24-hour Weighed Food Record is a validated method of assessing dietary intake. The Food Record has been adapted for New Zealand participants and reviewed by a registered dietician. For historical data records where a longer food record was administered, only the first day of the food record will be used in the analysis.

Participants will visit the Sensory Neuroscience Lab to receive their Food Record and instructions on how to complete the record. Participant responses will be transferred into daily energy intake (in kj) of different nutrient sources using FoodWorks (Xyris Software, Australia).

### 2.8 Secondary Outcomes

The following secondary outcomes will be assessed using the 24-hour Weighed Food Record:

- Fat intake
- Carbohydrate intake
- Protein intake
- Sugar intake
- Salt intake
- Micronutrients intake
- Animal protein
- Plant-based protein
- Alcohol consumption
- Fibre intake
- Healthy Eating Index

In addition to the Food Record, we will also collect the following measures:

- *Eating Behaviour:* Participants will be administered the Dutch Eating Behavior Questionnaire (DEBQ) (31) via online survey prior to collecting their Food Record. The DEBQ is a measure of individual eating behaviour consisting of 33 items on a 5-point scale (never to very often) that contribute to scales measuring emotional, external, and restrained eating.
- *BMI*: Height and weight will be collected during the study visit using a stadiometer and calibrated scales and used to calculate BMI.

### 2.9 Baseline

Baseline demographics will be pre-existing data of the pre-COVID cohort. Participants in the post-COVID infection cohort will complete an online survey which includes a baseline survey. This survey includes demographic information (year of birth, ethnicity, gender, socioeconomic) and experience with COVID-19 infection including timing, symptoms and severity, and experience of sensory dysfunction. Participants will also be asked about any changes they have perceived in their food choices and eating behaviour since before the pandemic.

## 3 Statistical analysis

Statistical analysis will be conducted using IBM SPSS Statistics 30.0 (IBM, Armonk, NY) and RStudio (PBC, Boston, MA). Baseline characteristics of participants will be summarised for the pre-COVID cohort and post-COVID cohort using mean and standard deviation (SD) for continuous variables and frequencies and percentages for categorical variables.

Total daily energy intake will be compared across cohorts using an ANCOVA analysis controlling for age, gender, and ethnicity. Subgroup analysis will be conducted for data collected before March 2020 and March 2020-July 2021 in order to analyse the specific effect of public health measures to control COVID in the absence of infection. Subgroup analyses will be conducted based on the dominant variant at the time of infection.

### 3.1 Missing data

Missing data will be reported but excluded from analysis. We will exclude participants from analysis that are missing their food record. Incomplete or implausible food records will be excluded.

## 4 Discussion

Understanding the impact of COVID-19 infection on long-term dietary choices and nutrition is crucial to ensure that public health measures are in place to respond to COVID-19, as well as future disease outbreaks. COVID-19 has had a disparate impact on different groups, with an increased burden on groups who already face unequal treatment and outcomes in the health system, such as Māori and Pacific Islanders, and individuals from more deprived neighbourhoods. Responding to COVID-19 and the potential impact on dietary choices is an opportunity to close the health gap and better respond to the health needs of vulnerable communities.

While COVID-related sensory changes are hypothesised to have impacts on eating behaviour and dietary choices, it is challenging to perform controlled cohort studies due to the high prevalence of undetected infections. The proposed temporal analyses provide a unique opportunity to test for COVID-related impacts on eating behaviour.

## Data Availability

All data produced in the present study are available upon reasonable request to the authors

